# Combined Use of Topic-Analysis and LLMs on Online Peer-to-Peer Medicaid Discussions to Detect the Impact of HR1 on Individuals’ Medicaid Experience Across The US

**DOI:** 10.64898/2026.11.03.26367093

**Authors:** Valmeek Kudesia

## Abstract

Medicaid is a government program providing benefits to 1 in 5 people in the US, including medical care for children and adults, long-term care for the elderly, and home/community-based services for the disabled[1]. Effective January 1st 2027, HR1 (“One Big Beautiful Bill”) passed by Congress will implement new administrative requirements to Medicaid expected to strip benefits from 1.8 million adults, children, and seniors[2]. As each US state administers Medicaid independently, the timing, frequency, and severity of those losses may vary across states. Monitoring that human impact requires an detailed ongoing view of affected individuals and their families. To monitor the human-impact of HR1, this work develops a proof-of-concept using topic analysis and LLMs to generate detailed categorization of individuals’ Medicaid experiences across multiple states and over time.

## Introduction

Medicaid is a government program providing benefits to 1 in 5 people in the US, including medical care for children and adults, long-term care for the elderly, and home/community-based services for the disabled[1]. Effective January 1st 2027, HR1 (“One Big Beautiful Bill”) passed by Congress will implement new administrative requirements to Medicaid expected to strip benefits from million adults, children, and seniors[2]. As each US state administers Medicaid independently, the timing, frequency, and severity of those losses may vary across states. Monitoring that human impact requires an detailed ongoing view of affected individuals and their families. To monitor the human-impact of HR1, this work develops a proof-of-concept for detailed categorization of individuals’ Medicaid experiences across multiple states and over time.

Since 2012, people across the US have posted Medicaid questions and experiences to a public peer-to-peer Medicaid-specific discussion forum. Large language-models (LLMs) can summarize and organize information from such text corpora but are limited by the amount of text consumable at a time i.e. “context window”. LLMs may miss patterns that unfold over time-segmented analyses or occur at frequency below a sampling threshold. Topic analysis is a complementary technique can efficiently analyze the entire corpus at once, however, its output are difficult to interpret. This work investigated the combined use of topic analysis with LLMs generate a readable categorization of individuals’ common and rare Medicaid experiences from 2012 until current day that is sufficiently detailed and interpretable to monitor the human impact of HR1.

## Methods

This work compared the performance of 1) 2 topic analysis techniques and 2) 5 LLMs to detect and summarize Medicaid-related themes of conversation (“topics”) contained in an online peer-to-peer public discussion group active for the past 14 years, Figure 1. A LLM only approach would only accommodate an estimated 4-6 months of conversations which would ignore conversations prior to HR1 requirements. To incorporate all conversations, an alternative approach was used that first performed topic analysis on all conversations over the entire time duration to detect topics and then provide those topics with representative conversations to LLMs for interpretation. The combination of topic analysis with LLMs is critical because topic analysis techniques are unsupervised and their outputs that represent a conversation theme, “topic names” may be difficult to interpret; however, a LLM provides readable interpretations of topic and topic names outputs. The “Turftopic” python library v0.17.2 was used for topic analysis and temporal fitting [3].National and state-level Medicaid enrollment[4] and program characteristics[5] were included in comparisons. All computation was performed on modest local hardware, refer to Appendix for details.

**Figure 1:**
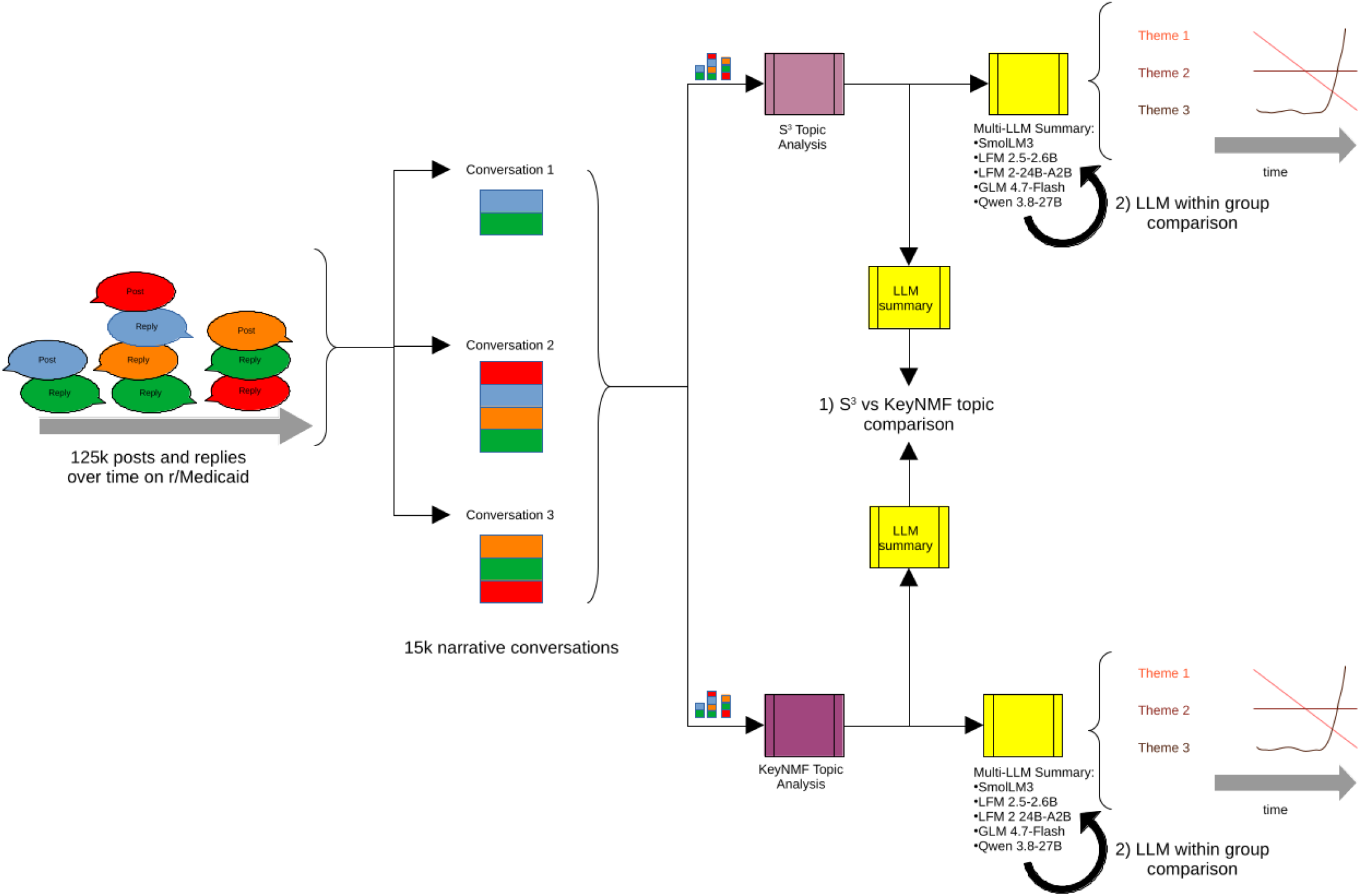
Summary of methods that generate corpus for analysis, deploy topic analysis and LLMs, and comparisons between 1) topic analysis techniques and 2) LLMs

### Corpus

The online conversations included public peer-to-peer discussions on www.reddit.com (r/Medicaid) over the time period of 02/01/2012-07/01/2026, data retrieved 7/12/2016. Each online post and related replies were assembled into a single narrative conversation. resulting in a corpus of 15,170 conversations with associated date and time, Figure 1. The 15,170 conversations totaled 38.6M characters which vastly exceeded the 1M context window of most available LLMs

### Comparison of Topic Analysis Techniques

Two dynamic topic analysis methods separately analyzed the entire corpus of 15,170 conversations to capture the time course of 100 distinct topics over the study period: Semantic Signal Separation (S^3^) and KeyNMF. S^3^ is a newer semantic content-based approach capable of detecting subtle nuance between conversation themes, while KeyNMF is a more traditional entity-and-keyword approach[3]. The count of 100 topics for 15,170 conversations was chosen to standardize comparisons and maintain an average ratio of about 1 topic per 100 conversations. The ratio of about 1 topic per 100 conversations is a very conservative limit that ensures the topic analysis techniques will identify topics with abundant evidence over the entire time duration in the corpus. Topics were compared against each other via summary from a single LLM with instruction to limit output to about 5 words. See Figure 1 “1) S^3^ vs KeyNMF comparison.”

### Comparison of LLM Performance

Each conversation theme over time produced by S^3^ and KeyNMF along with the 10 most representative conversations for that theme were each summarized by the same 5 open-source LLMs at Q4 quantization: SmolLM3[6], LFM 2.5-2.6B[7], LFM 2-24B-A2B[7], GLM 4.7-Flash[7], and Qwen 3.8-27B[8], see Figure 1 “2) LLM within group comparison.” Open source LLMs were chosen as they provide the most transparency into model performance. Specific LLMs were selected to represent a range of capability and sophistication. No tailored prompt engineering was applied; the same simple instruction was provided to each LLM. All parameters, including “thinking”, were set to default.

## Results

### Corpus

The study corpus included 125,332 public discussion posts by 21,145 users which totaled 38,603,217 characters. Discussion posts spanned from the creation of the Medicaid forum (02/01/2012) until date data obtained (07/01/2026). Over that time period the number of discussion posts regarding Medicaid increased 100-fold: 10 posts in 2012 vs 16,162 posts in 2026 whereas Medicaid enrollment increased 2-fold over the same time period[4]. Medicaid-focused conversations on r/Medicaid are granular to the level of the individual’s experience, refer to Appendix for examples.

### Comparison of Topic Analysis Techniques

The following sections correspond to “1) S3 vs KeyNMF topic comparison” in Figure 1.

### S^3^ Topic Analysis

The 100 topics detected by S^3^ address Medicaid eligibility, enrollment, administrative processes, and coverage across single or multiple states with process-specific and state-specific nuance e.g. “how to resolve an incorrect denial” or “household definition in North Carolina” and show good separation over time. The top 5 topics over the study duration addressed Medicaid applications, transitioning from children’s to adult Medicaid, income calculations, provider coverage, and billing disputes, Figure 2. The 5 topics with the most increase of importance addressed work waivers, reaching customer service and high risk pregnancies, unreliable Medicaid application technology, and issues related to income, Figure 3. Sixteen (16) topics specifically addressed an impact to family, children, or elders. Twenty-five states (25) were specifically mentioned across 19 topics, either alone or as a group. State Medicaid enrollment over time did not appear correlated with the 19 topics’ importance over time. Of the 25 states, 21 have implemented Medicaid expansion, 24 utilize Medicaid Managed Care Organizations (MCOs), and 5 utilize primary care case management (PCCM). The top 5 topics over study duration that specifically reference a state addressed a similar range of concerns. Refer to Appendix for heatmap of top 5 state-specific topics and all 100 topics.

**Figure 2:**
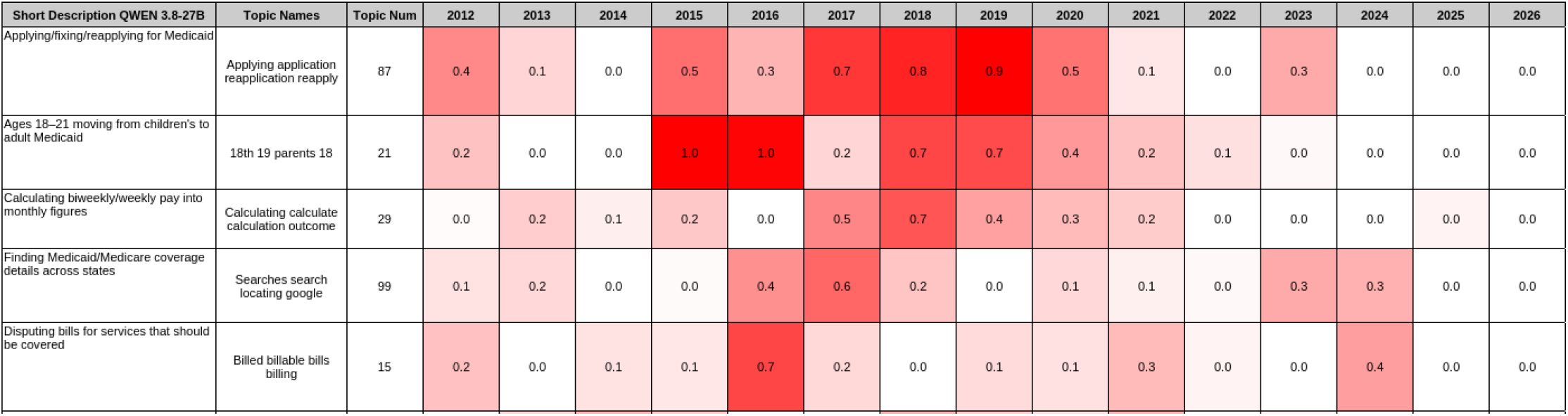
S^3^ 5 most-important topics over study duration. Darker = more importance. Values are dimension-less

**Figure 3:**
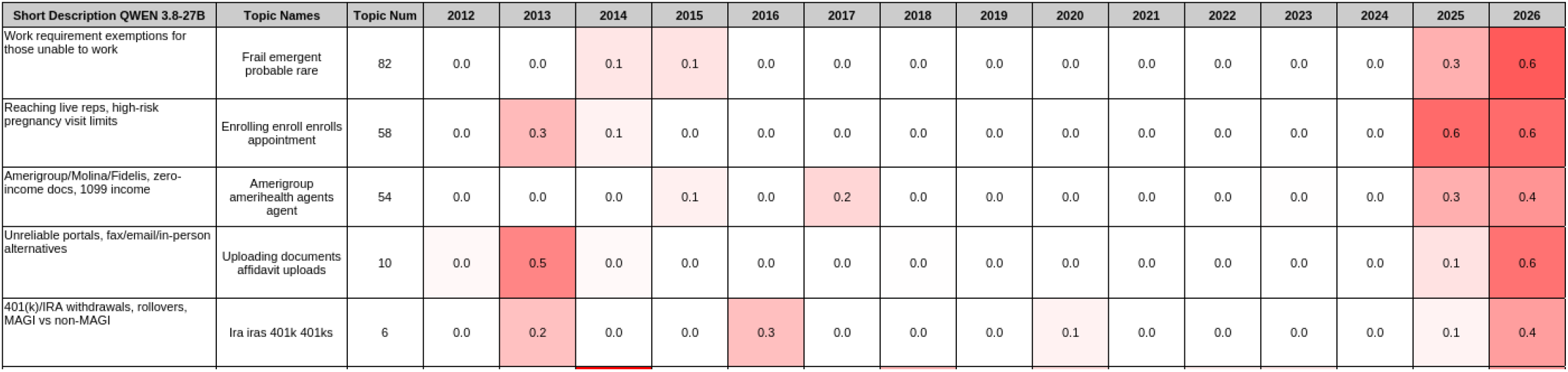
S^3^ 5 topics with most increased importance over study duration. Darker = more importance. Values are dimensionless

### KeyNMF Topic Analysis

KeyNMF topics focused on policy or organizational issues e.g. political advocacy or healthcare premiums and a single topic dominated throughout the duration of the study. Overall, KeyNMF topic importance was found to be more homogeneous over time and concentrated in few topics which is not suited for this application. Refer to Appendix for KeyNMF topic heatmaps.

### Comparison of LLM Performance within Single Topic Analysis Technique

This section corresponds to “2) LLM within group comparison” in Figure 1. The 5 different LLMs were each separately provided the topic names with 10 most representative conversations from S^3^ simply tasked to “interpret the topic” without tailored prompt engineering or specific word limit. Topics generated via KeyNMF were not used in the LLM comparison as planned given the discovered limitations of KeyNMF topics. On average, LLMs with more parameters provided more efficient topic summarization whereas LLMs with fewer parameters produced more verbose summarization: GLM-4.7 Flash 30B-A2.6B (384 characters), QWEN-3.8 27B (526 characters), LFM2-24B-A2B (626 characters), SmolLM3 (1652characters), LFM-2.5 2.6B (630 characters). LFM-2.5 2.6B was the exception with had the least number of parameters (2.6B) but produced relatively concise summaries given its very low number of parameters, see Table 1. Refer to Appendix for sample of multi-LLM summarization of S^3^ topics.

**Table 1.** Sample LLM summary output for S^3^ topic with most increase in importance over time. LLMs are arranged in order of most parameters (30B) at the top to least parameters (2.5B) at the bottom. <u>Topic names</u>: Frail emergent probable rare <u>Short description</u>: Work requirement exemptions for those unable to work

|  |  |
|---|---|
| GLM-4.7 Flash 30B-A2.6B | The topic describes discussions regarding the eligibility and coverage specifics of Medicaid, including federal poverty levels, work requirements, medically frail status exemptions, and retroactive coverage gaps. |
| QWEN-3.8 27B | This topic centers on the challenges faced by individuals navigating Medicaid eligibility, particularly those who are medically frail or unable to work. It covers how work requirements introduced under federal and state legislation affect beneficiaries, the process of determining and renewing medically frail status, and the financial and bureaucratic hurdles people encounter when transitioning between coverage, dealing with gaps in insurance, or trying to maintain eligibility while managing serious health conditions. |
| SmolLM3 3B | Based on the provided information, the topic revolves around the discussion of federal Medicaid work requirements, Medicaid coverage for braces, and questions regarding the retroactive coverage of medical expenses and the process of transitioning from one insurance plan to another. The main concerns include:<br>1. Determining whether Medicaid covers braces again after a lapse in coverage.<br>2. Understanding the retroactive coverage of medical expenses when switching insurance plans.<br>3. The process of switching from one health insurance provider to another and the potential impact on medical expenses.<br>4. The role of Medicaid in covering medical expenses when switching between insurance plans.<br>5. The availability of financial aid or charity care for individuals who may not be covered by their new insurance plan.<br>The thread also touches on various states' Medicaid work requirements, specifically Nebraska and Indiana, and the process of determining medical frailty status. Additionally, there are questions about the applicability of medical care for specific conditions, such as migraines and PTSD, and the eligibility for Medicaid benefits.<br>It seems that the participants are seeking advice on the following:<br>• Whether their past medical conditions or procedures, such as braces or migraines, will be covered under a new insurance plan.<br>• The process of applying for and transitioning between insurance plans and the potential impact on their medical coverage.<br>• The retroactive coverage of medical expenses when switching insurance providers.<br>• The eligibility for financial aid or charity care when the new insurance plan does not cover certain medical expenses.<br>• The specifics of state Medicaid work requirements and the process of determining medical frailty status.<br>• The availability of specific medical services, such as braces or mental health care, and the eligibility criteria for these services. |
| LFM-2.5 2.6B | This topic involves individuals dealing with Medicaid eligibility, work requirements, and medical insurance challenges, particularly focusing on those who are medically frail or have rare conditions. Several users share experiences with navigating work requirements that were introduced in 2023, especially in states like Indiana and Nebraska, where medically frail status is under review more frequently. One user discusses the transition to Medicaid buy-in and the use of third-party trusts to save for future needs. Another user from Nebraska describes the difficulties of securing health insurance and meeting work requirements as their eligibility period changes. Several threads also address questions about whether Medicaid can cover specific expenses like braces or whether having gaps in coverage can break the law, highlighting the complexity and variability of state Medicaid programs. Overall, the content reflects a group of people seeking clarity and assistance with navigating the intricate rules and requirements of Medicaid and related health insurance coverage. |

## Discussion

Topic modeling by S^3^ is sufficiently granular to capture concerns related to Medicaid enrollment, eligibility, work requirements, and administrative obstacles, generally and related to specific states. S^3^ topics related to unexpected billing and provider network coverage match to primary survey findings[10]. In addition, S^3^ readily identifies topics with relation to family, children, and elderly. This is especially important given Medicaid’s importance to pediatric and eldercare in the US. In light of the upcoming HR1 changes to Medicaid and state-by-state implementation, this proof-of-concept demonstrates an ability to observe at least some of the ongoing effects on millions of individuals’ state-level Medicaid experience. For example, as work-requirements are implemented we may see increased peer-to-peer conversations describing confusion regarding the process of attesting to work-requirements in each state, particularly if many individuals are employed via the gig-economy. In another example, we may see increased peer-to-peer conversation describing older individuals who have been administratively disenrolled from Medicaid and now face difficulty in paying for long-term care. The grouping of multiple US states via S^3^ topics (e.g. S^3^ topic 21 which groups NC, NE, NY) is unexpected and possibly suggests some underlying similarity in the individuals’ granular Medicaid experiences in those states. LLMs with parameter count >=27B appear better suited for this summarization task; however, LFM-2.5 2.6B had the fewest parameters and lowest computation costs yet provided fairly compact topic descriptions thereby making it the most efficient LLM for this task. This appears consistent with authors’ assertion of uniquely efficient model architecture. LFM-2.5-2.6B appears well suited for ongoing efficient re-summarization of topics from ongoing conversations to monitor for established or emerging effects from HR1.

This proof-of-concept has significant limitations. The peer-to-peer discussions are primarily in English and populations centers are likely over-represented. The anonymous nature of this online discussion encourages questions and commentary yet limits the ability to verify specific events or experiences. The conservative 1:100 ratio for topic detection means some medium to low frequency topics in the corpus were ignored by the topic analysis techniques, which is acceptable for this proof-of-concept. Future work will include HR1 effects on Supplemental Nutrition Assistance Program (SNAP), Temporary Assistance for Needy Families (TANF), and other governmental benefits.

## Data Availability

All data produced in the present study are available upon reasonable request to the authors

## Appendix

**Table 2.**
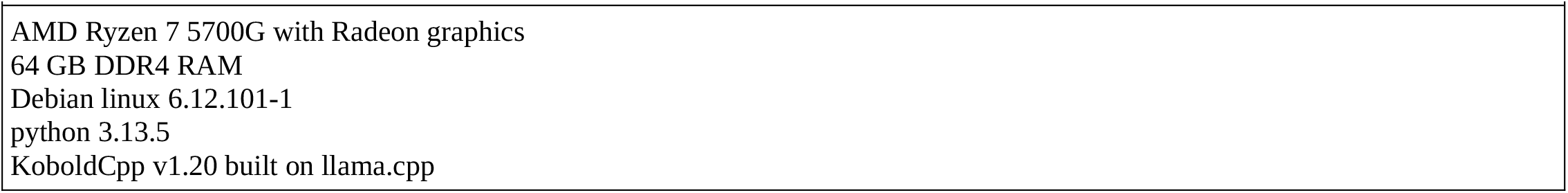
Computational details.

|  |
|---|
| AMD Ryzen 7 5700G with Radeon graphics<br>64 GB DDR4 RAM<br>Debian linux 6.12.101-1<br>python 3.13.5<br>KoboldCpp v1.20 built on llama.cpp |

**Table 3.**
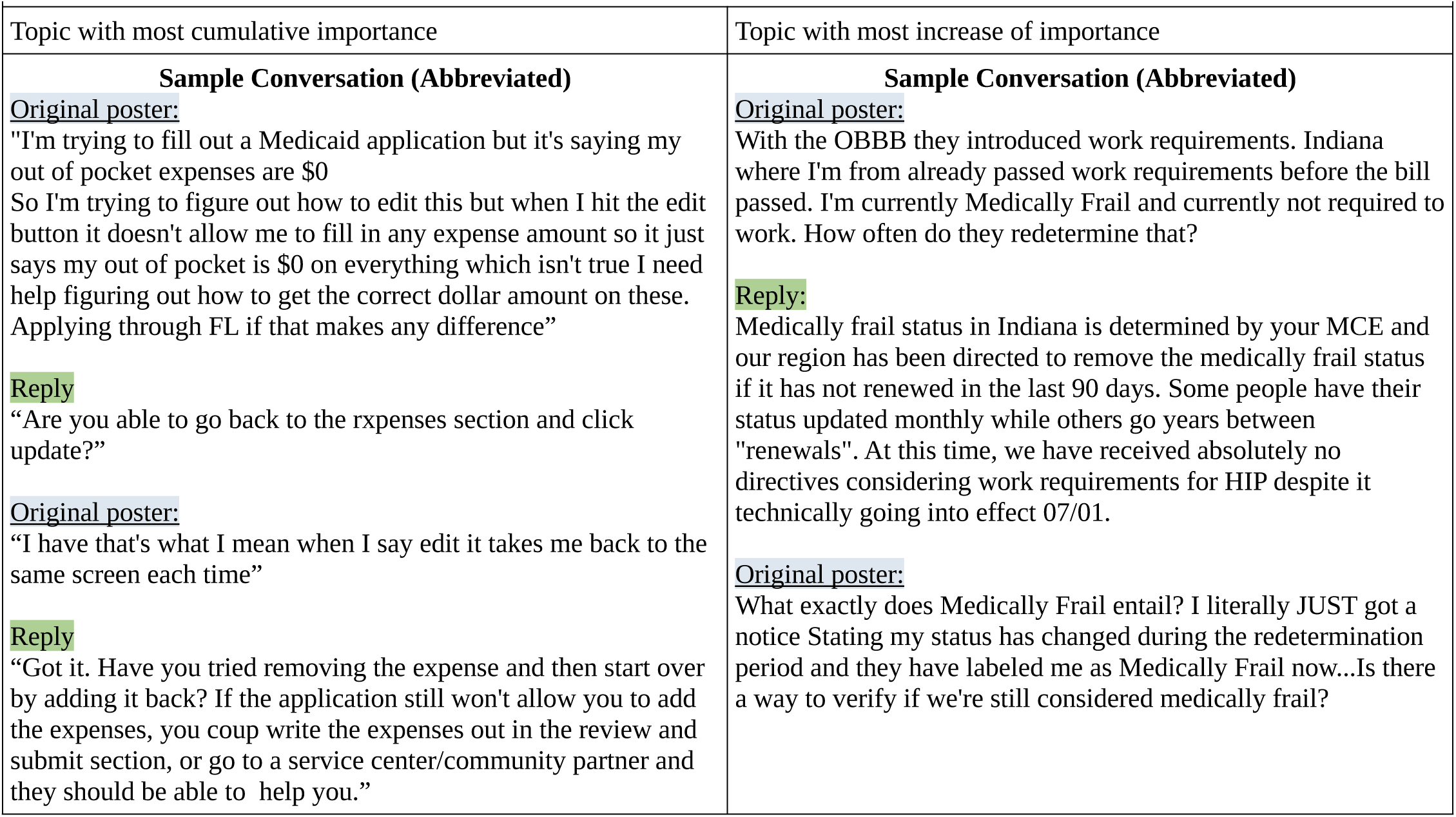
Sample conversation S^3^ topics.

| Topic with most cumulative importance | Topic with most increase of importance |
|---|---|
| **Sample Conversation (Abbreviated)**<br>Original poster:<br>"I'm trying to fill out a Medicaid application but it's saying my out of pocket expenses are \$0<br>So I'm trying to figure out how to edit this but when I hit the edit button it doesn't allow me to fill in any expense amount so it just says my out of pocket is \$0 on everything which isn't true I need help figuring out how to get the correct dollar amount on these.<br>Applying through FL if that makes any difference”<br>Reply<br>“Are you able to go back to the rxpenses section and click update?”<br>Original poster:<br>“I have that's what I mean when I say edit it takes me back to the same screen each time”<br>Reply<br>“Got it. Have you tried removing the expense and then start over by adding it back? If the application still won't allow you to add the expenses, you coup write the expenses out in the review and submit section, or go to a service center/community partner and they should be able to help you.” | **Sample Conversation (Abbreviated)**<br>Original poster:<br>With the OBBB they introduced work requirements. Indiana where I'm from already passed work requirements before the bill passed. I'm currently Medically Frail and currently not required to work. How often do they redetermine that?<br>Reply:<br>Medically frail status in Indiana is determined by your MCE and our region has been directed to remove the medically frail status if it has not renewed in the last 90 days. Some people have their status updated monthly while others go years between "renewals". At this time, we have received absolutely no directives considering work requirements for HIP despite it technically going into effect 07/01.<br>Original poster:<br>What exactly does Medically Frail entail? I literally JUST got a notice Stating my status has changed during the redetermination period and they have labeled me as Medically Frail now...Is there a way to verify if we're still considered medically frail? |

**Figure 4:**
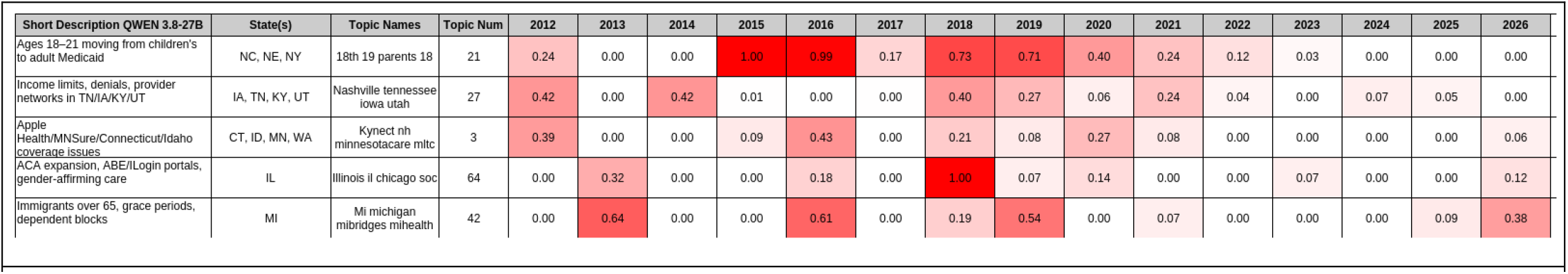
S^3^ 5 most importance topics specific to state over study duration. Darker = more importance. Values are dimensionless

**Figure 5:**
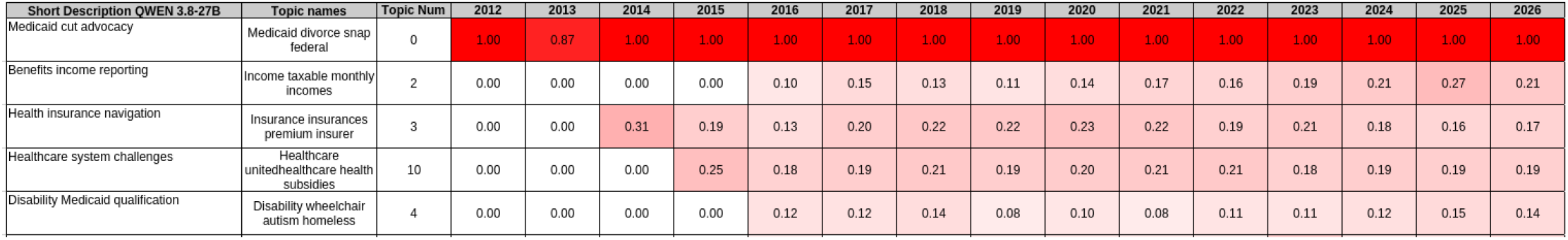
KeyNMF most important topics over study duration. Darker = more importance. Values are dimension-less

**Figure 6:**
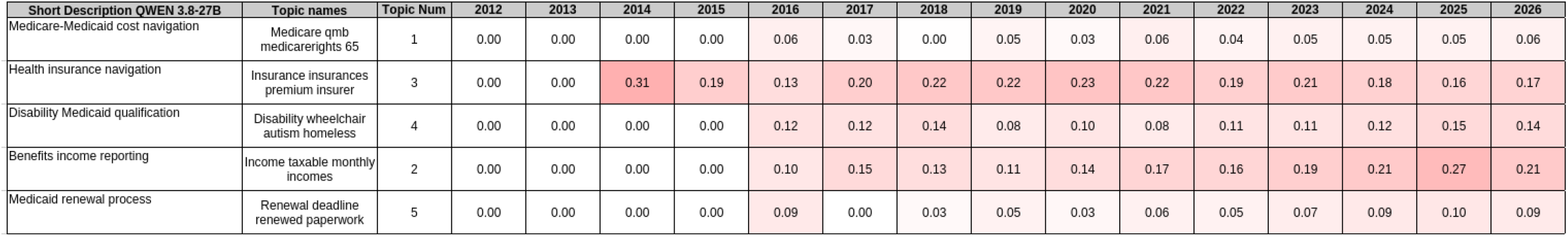
KeyNMF topics with most increased importance over study duration. Darker = more importance. Values are dimensionless

**Figure 7:**
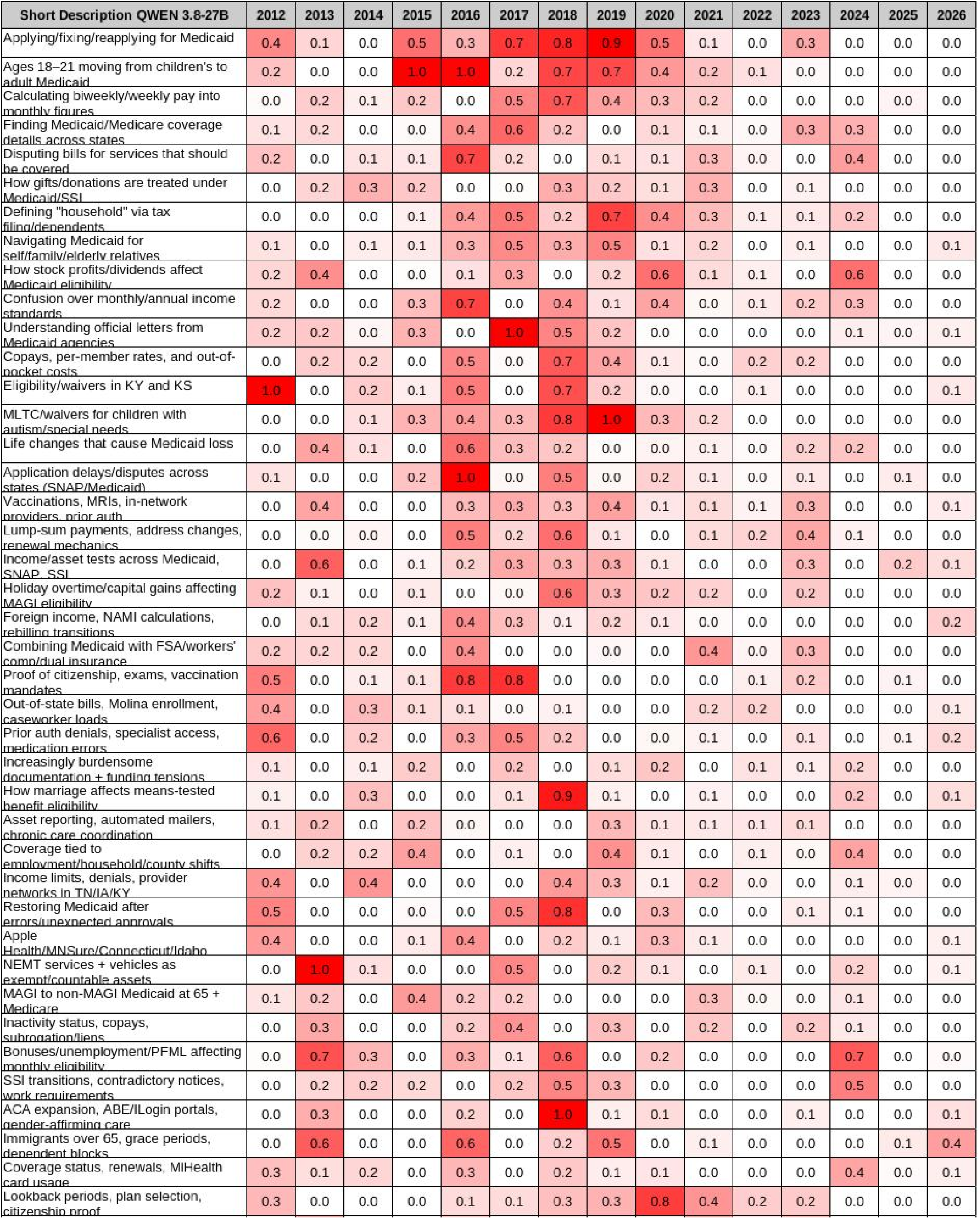
S3 topics with short description and importance over time. Arranged in descending importance over all time. Heat-map continued across multiple pages. Darker color implies more importance. Heat-map values are relative dimension-less.

**Figure 8:**
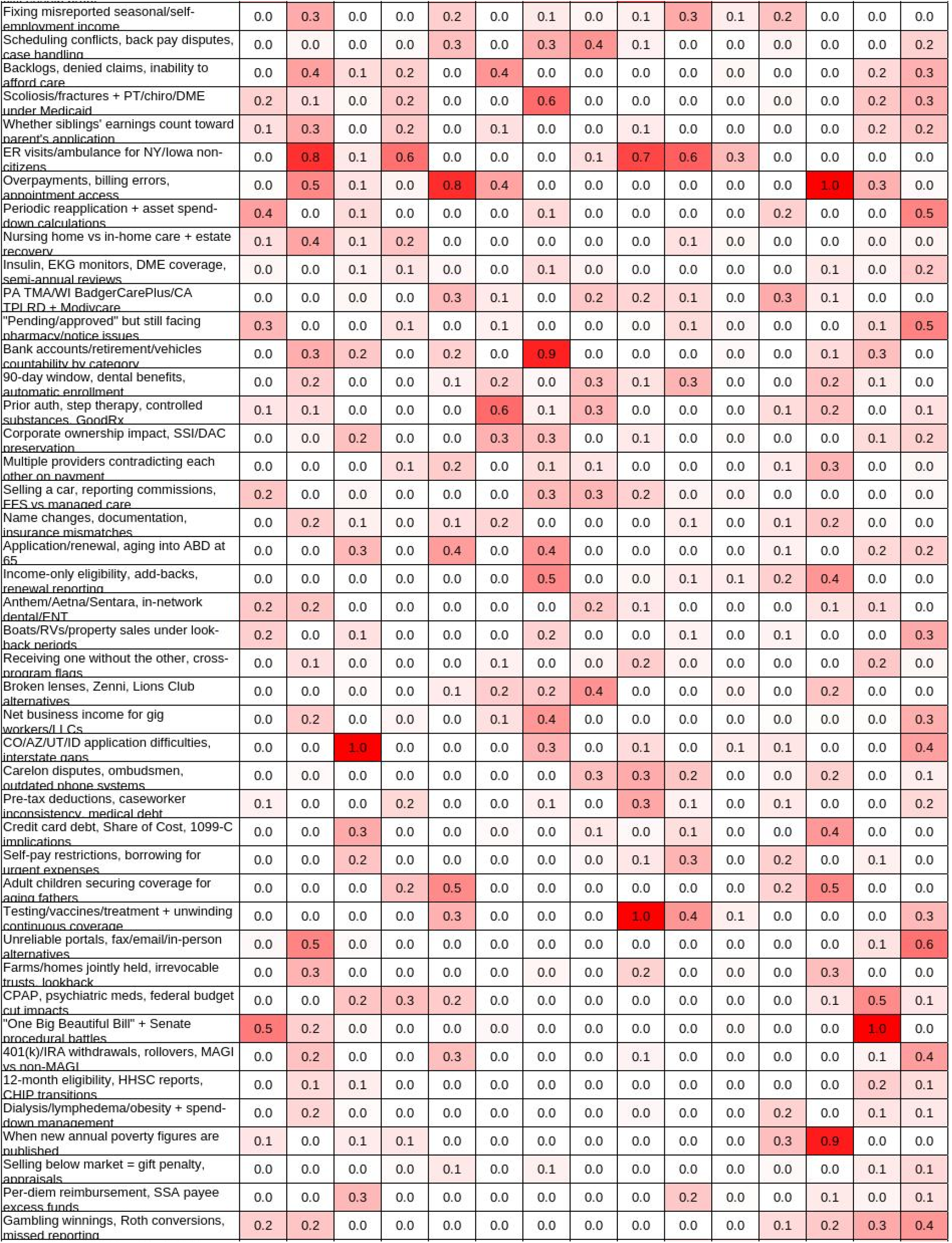
S3 topics with short description and importance over time. Arranged in descending importance over all time. Heat-map continued across multiple pages. Darker color implies more importance. Heat-map values are relative dimension-less.

**Figure 9:**
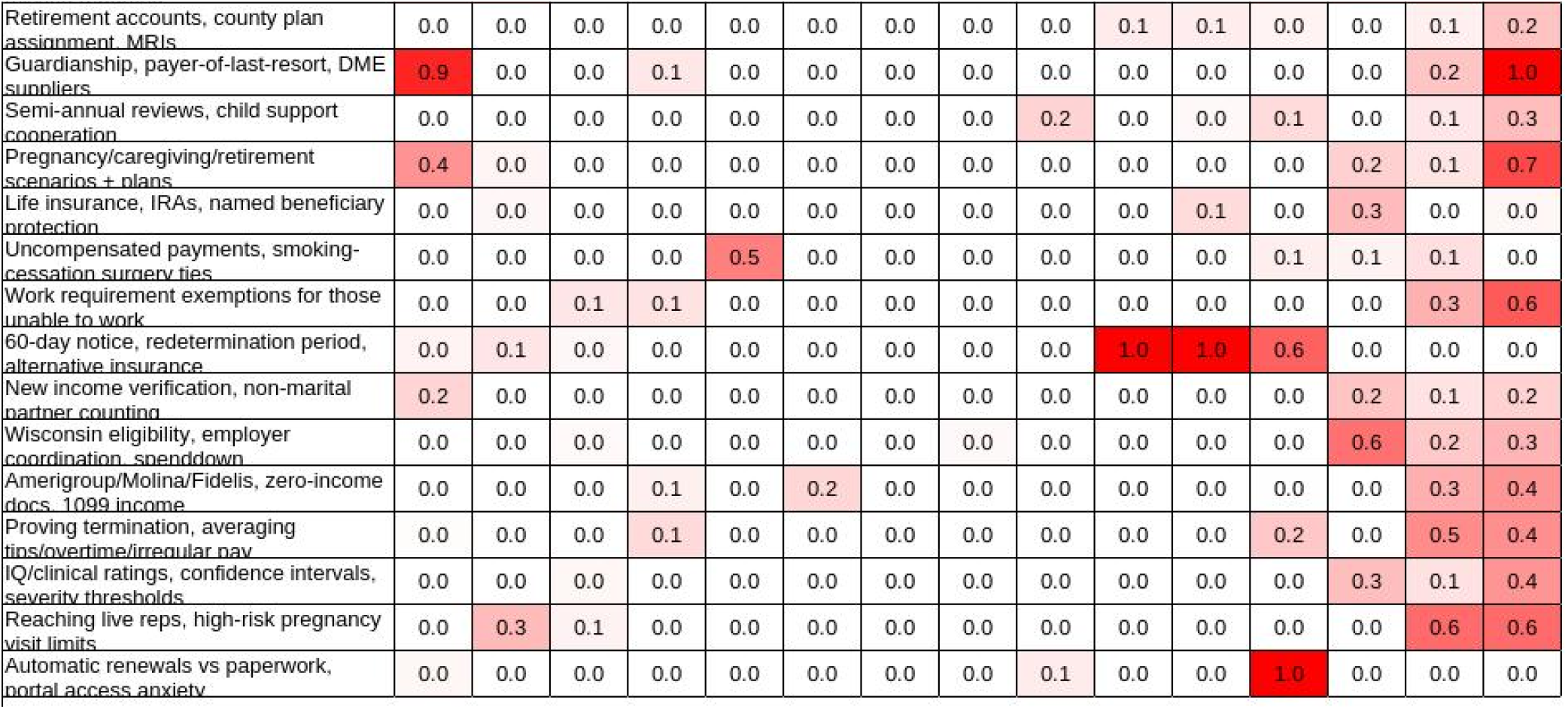
S3 topics with short description and importance over time. Arranged in descending importance over all time. Heat-map continued across multiple pages. Darker color implies more importance. Heat-map values are relative dimension-less.

**Figure 10.**
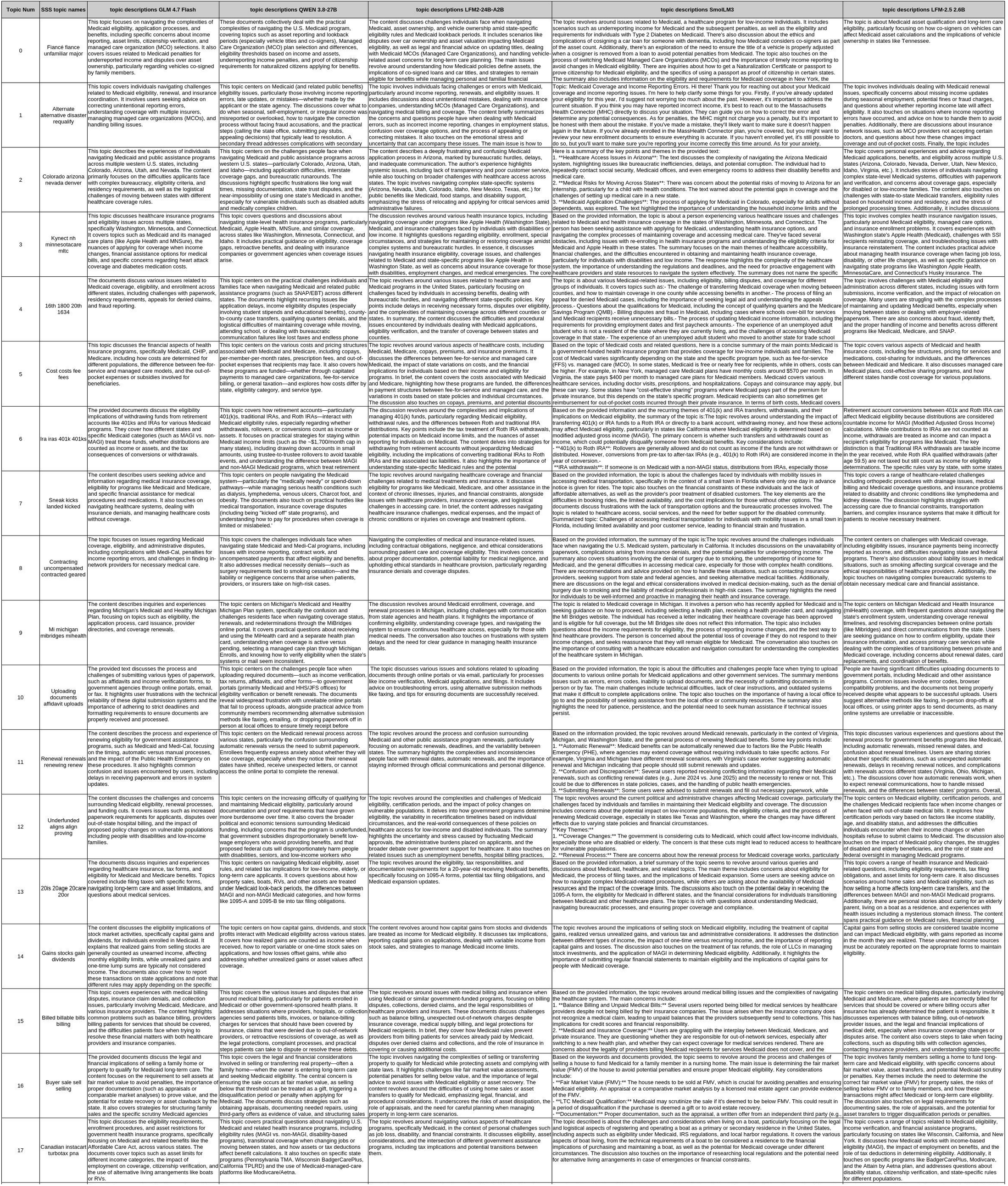
Multi-LLM descriptions of S^3^ topics. LLMs arranged from greatest (L) to least (R) number of parameters.

